# Vaccination and COVID-19 dynamics in hemodialysis patients: a population-based study in France

**DOI:** 10.1101/2021.07.06.21259955

**Authors:** Khalil El Karoui, Maryvonne Hourmant, Carole Ayav, François Glowacki, Cécile Couchoud, Nathanaël Lapidus, On Behalf Of The Rein Registry

## Abstract

**Importance:** Maintenance hemodialysis (MHD) patients have a high mortality risk after COVID-19 and an altered humoral response to vaccines, but vaccine clinical efficacy remains unknown in this population.

**Objective:** To estimate the association between vaccination and COVID-19 hospitalization rate in MHD patients

**Design:** Using Bayesian multivariable spatiotemporal models, we estimated the expected number of SARS-CoV-2 severe infections (infections with hospital admission) in MHD patients from simultaneous cases in the general population.

**Setting:** French population-based retrospective analysis in MHD and non-dialysis patients.

**Participants:** Models were fitted from 3620 hospitalizations of MHD patients and 457,160 hospitalizations in the general population.

**Exposure:** Severe SARS-CoV-2 infections in the general population and vaccine exposure.

**Main Outcome and Measure:** Weekly incidence of severe infections in MHD patients.

**Results:** During the first epidemic wave, incidence of severe infections in MHD patients was approximately proportional to incidence in the general population. However, our model overestimated incidence during the second wave, suggesting an effect of prevention measures during the 2^nd^ wave. A second model (based on data up to the end of the 2^nd^ wave) estimated that the risk in MHD patients decreased between waves 1 and 2, with incidence rate ratio (IRR) = 0.70 (95% CI: 0.64, 0.76). Moreover, while this model correctly estimated the reported MHD cases up to the end of the 2^nd^ wave, predictions overestimated the expected number of cases from the beginning of the vaccination campaign. Using vaccination coverages as additional predictors permitted to correctly fit the weekly reported number of cases, with IRR in MHD patients of 0.41 (95% CI: 0.28, 0.58) for vaccine exposure in MHD patients and 0.50 (95% CI: 0.40, 0.61) per 10% increase in vaccination coverage in the same-age general population.

**Conclusions and Relevance:** Our findings suggest that both individual and herd immunity due to vaccination may yield a protective effect against severe forms of COVID-19 in MHD patients.

**Question:** Whether vaccination against SARS-CoV-2 limits hospitalization rates in hemodialysis patients is still unknown.

**Findings:** By modeling the dynamics of 3620 hospital admissions for SARS-CoV-2 infections among hemodialysis patients, as a proportion of 457,160 cases reported in the French general population from March 2020 to April 2021, we identified vaccination coverage in both hemodialysis patients and the general population as independently associated with protection of hemodialysis patients against severe infection.

**Meaning:** Vaccination against SARS-CoV-2 is associated with reduced hospitalization rate in hemodialysis patients.

## Introduction

Patients with end stage renal disease requiring maintenance hemodialysis (MHD) have a 20-25% risk of mortality after infection with Severe Acute Respiratory Syndrome Coronavirus 2 (SARS-CoV-2)^1–3^. Moreover, altered humoral responses after SARS-CoV-2 vaccination have been described in MHD population^4^. These results led to prioritize vaccination in MHD patients in France in early 2021, leading to a high vaccination coverage compared to the non-dialysis population. However, the evaluation of vaccine efficacy against severe forms of COVID-19 in this population remains an unmet need.

## Methods

We analyzed aggregated weekly counts of severe SARS-CoV-2 infections (infections followed by hospital admissions) in MHD patients and in the French general population. Cases in MHD patients were reported within the French end-stage renal disease REIN registry from March 11, 2020 to April 29, 2021^2,5^. Vaccine exposure in MHD patients (first dose) was reported in weekly surveys sent to all French nephrology centers since January 2021.

Weekly incidences of severe infections in the general population were obtained from the national exhaustive inpatient surveillance system (SI-VIC database), assuming a 11-day time lapse from infection to hospitalization^6^. Weekly vaccination coverage (first dose) in the general population was obtained from the National surveillance system (VAC-SI database)^7^.

We estimated the expected weekly incidence of severe infections in MHD patients from simultaneous cases reported for the same age class in the general population, using Bayesian hierarchical Poisson regressions with the log number of at-risk MHD patients as an offset. Model M_1_ only used incident cases from the first epidemic wave (up to June 30, 2020). Model M_2_ used similar data up to the end of the second wave and beginning of the vaccination campaign in MHD patients (January 7, 2021), considering a possible risk change between the two waves. Model M_3_ used similar data up to April 29, 2021 with vaccine coverage in the general population and in MHD patients as additional predictors. Weekly vaccination coverage data were lagged by 3 and 5 weeks in the general population and MHD patients, respectively, to account for a possible effect of constituted immunity on infection risk^4,8–10^. Only subjects aged at least 25 years were considered from all data sources. Statistical analyses are detailed in supplementary material.

## Results

Vaccination coverage in MHD and the general population is shown in Figure 1A. Approximately 45,000 MHD patients were at-risk during the follow-up, among whom 3620 SARS-CoV-2 severe infections were reported (Figure 1B). In the meantime, 457,160 hospital admissions were reported in the general population (Figure 1B). Model M_1_ showed that during the first wave, incidence in MHD patients was approximately proportional to incidence in the general population (Figure 1C, Supplementary Table S1). Incidence rate ratios (IRR) were higher in younger MHD patients: compared to patients aged 55-65 years, IRR were 4.04 (95% credible interval [CI]: 2.38, 6.51) and 0.87 (95% CI: 0.73, 1.04) in patients aged 25-35 and 75-85 years, respectively, suggesting that younger MHD patients have a relatively higher risk of hospitalization than older MHD patients, when compared to the same-age general population (Supplementary Table S2).

**Figure 1:**
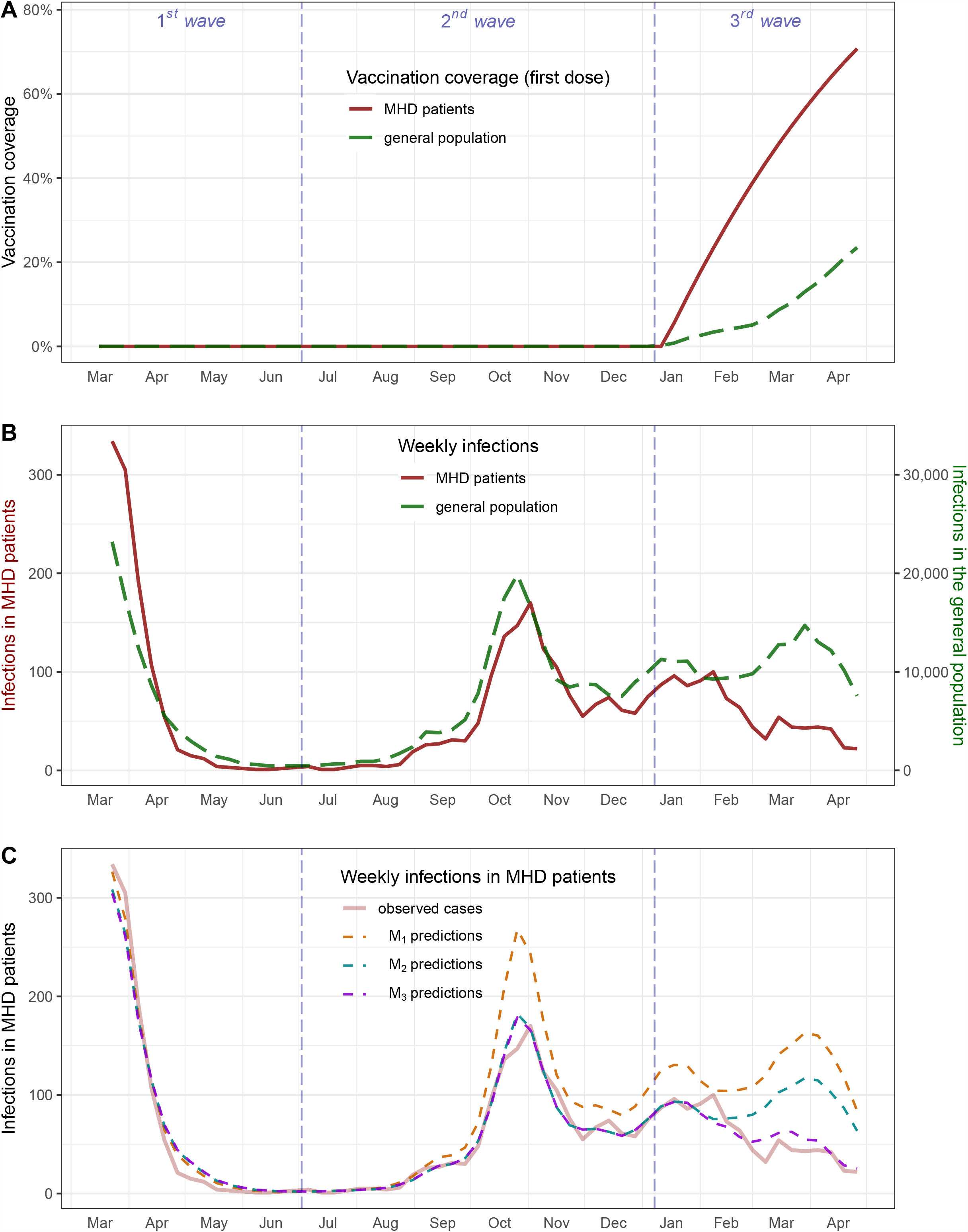
Nationwide vaccination coverage, and SARS-CoV-2 severe infections in patients needing maintenance hemodialysis (MHD) and in the general population (adults aged at least 25 years). **Panel 1A:** vaccine policies started in mid-January and first targeted patients with compromising health conditions such as MHD patients among whom vaccination quickly reached a high coverage. **Panel 1B:** incidence of severe infections in MHD patients approximately followed the dynamics in the general population during the two first epidemic waves. The relative incidence in MHD patients dropped in early 2021, following the beginning of the vaccine policies. **Panel 1C:** Model M_1_ fitted on 1^st^ wave data failed to capture an incidence drop in MHD patient incidence from the beginning of the second wave. Model M_2_ fitted on 1^st^ and 2^nd^ wave data failed to capture the early 2021 dynamics of MHD incidence, contrary to Model M_3_ using vaccine exposures as additional predictors.

Predictions from this model overestimated incidence during the second wave: 2092 predicted cases (95% CI: 1888, 2317) vs. 1447 reportedly (Figure 1C, Supplementary Table S1), suggesting an effect of prevention steps taken during the 2^nd^ wave.

Model M_2_ (data up to the end of the 2^nd^ wave) estimated that the risk in MHD patients decreased between waves 1 and 2, with IRR = 0.70 (95% CI: 0.64, 0.76). Moreover, while M_2_ correctly estimated the reported MHD cases up to the end of the second wave (Figure 1C), predictions from this model overestimated the expected number of cases from the beginning of the vaccination campaign (3^rd^ wave): 1457 (95% CI: 1356, 1557) vs. 940 reportedly (Figure 1C and Supplementary Table S1).

Using vaccination coverages as additional predictors in M_3_ permitted to correctly fit the weekly reported number of cases (Figure 1C). IRR were 0.41 (95% CI: 0.28, 0.58) for vaccine exposure in MHD patients and 0.50 (95% CI: 0.40, 0.61) per 10% increase in vaccination coverage in the same-age general population, suggesting that vaccine exposure in both MHD patients and the general population is independently associated with reduction of hospitalization rate of MHD patients, when compared to the same-age general population (Supplementary Table S2). Coefficients from these models are reported in Supplementary Table S2.

## Discussion

By modeling the dynamics of SARS-CoV-2 infections leading to hospital admissions among MHD patients in France as a proportion of cases reported in the general population, we identified a relative reduction of observed cases in the 2^nd^ wave, compared to a model based on the 1^st^ wave only. This result suggests that, compared to the general population, the hospitalization rate of MHD patients in the 2^nd^ wave was lower than expected from data of the first wave. This observation may be explained by the progressive implementation of prevention steps, earlier testing and better management of COVID-19 of MHD patients during the 2^nd^ wave. Indeed, since the beginning of the epidemic, protective strategies have been broadcast by the French Society of Nephrology with weekly COVID-19 webinars. However, this effect could be also confounded by a trend to limit hospitalization of MHD patients with milder cases.

Moreover, our models also suggest a reduction of hospitalized cases starting in early 2021, compared to a model based on the 1^st^ and 2^nd^ waves only. Therefore, we hypothesized that vaccination exposure could have had a role in limiting hospital admissions in MHD patients. We took advantage of the French vaccine policies prioritizing MHD patients (Figure 1A). Taking into account vaccine exposure in both the general population and MHD patients, we (i) could adequately predict the observed incidence during the 3^rd^ wave, and (ii) found that the reduction of cases was independently associated with vaccine exposure in both MHD patients and the same-age general population. These findings suggest that vaccination may have a protective effect from the first dose in MHD patients and that these patients may indirectly benefit from vaccine policies at the population scale, probably thanks to a lessened exposure to the virus from their encounters. Importantly, vaccine exposure data were dependent of weekly reports from dialysis centers. Vaccine exposure could therefore be overestimated, since dialysis centers with high vaccination rates might tend to better report their data. Consequently, vaccine exposure appears as a major factor associated with reduction of hospitalization of MHD patients in early 2021.

The strength of our analysis is the large number of patients followed in the MHD population since the beginning of the SARS-CoV-2 spread in France, thanks to the nationwide REIN registry^2,5^. However, in absence of individual data on vaccination or previous SARS-CoV-2 infection, we were not able to evaluate the individual risk of severe form of COVID-19 post-vaccination^11^. Besides, whether this analysis could be extended to other populations with different prevention measures (including types of vaccines and vaccination schemes) remains to be studied. Of note, our analyses end before the strategy of third vaccine injection starts in France (in late April 2021). Lastly, our data did not include important virological factors such as the spread of variant types in both populations. However, no evidence to date suggests that SARS-CoV-2 variants could be different in MHD and non-dialysis patient infections.

In conclusion, this study identifies vaccination coverage in both MHD patients and the general population as independently associated with protection against severe infection. Though causal relationships cannot be demonstrated from such an observational study, our findings suggest that both individual and herd vaccine-induced immunity may yield a protective effect against severe forms of COVID-19 in MHD patients.

## Data Availability

Data are available upon request

## References

1. Williamson EJ, Walker AJ, Bhaskaran K, et al. Factors associated with COVID-19-related death using OpenSAFELY. Nature. 2020;584(7821):430–436. doi:10.1038/s41586-020-2521-4

2. Couchoud C, Bayer F, Ayav C, et al. Low incidence of SARS-CoV-2, risk factors of mortality and the course of illness in the French national cohort of dialysis patients. Kidney Int. 2020;98(6):1519–1529. doi:10.1016/j.kint.2020.07.042

3. Jager KJ, Kramer A, Chesnaye NC, et al. Results from the ERA-EDTA Registry indicate a high mortality due to COVID-19 in dialysis patients and kidney transplant recipients across Europe. Kidney Int. 2020;98(6):1540–1548. doi:10.1016/j.kint.2020.09.006

4. Ikizler TA, Coates PT, Rovin BH, Ronco P. Immune response to SARS-CoV-2 infection and vaccination in patients receiving kidney replacement therapy. Kidney Int. 2021;99(6):1275–1279. doi:10.1016/j.kint.2021.04.007

5. Couchoud C, Stengel B, Landais P, et al. The renal epidemiology and information network (REIN): a new registry for end-stage renal disease in France. Nephrol Dial Transplant Off Publ Eur Dial Transpl Assoc - Eur Ren Assoc. 2006;21(2):411–418. doi:10.1093/ndt/gfi198

6. Hozé N, Paireau J, Lapidus N, et al. Monitoring the proportion of the population infected by SARS-CoV-2 using age-stratified hospitalisation and serological data: a modelling study. Lancet Public Health. 2021;6(6):e408–e415. doi:10.1016/S2468-2667(21)00064-5

7. Données relatives aux personnes vaccinées contre la Covid-19 (VAC-SI) - data.gouv.fr. Accessed July 1, 2021. https://www.data.gouv.fr/en/datasets/donnees-relatives-aux-personnes-vaccinees-contre-la-covid-19-1/

8. Haas EJ, Angulo FJ, McLaughlin JM, et al. Impact and effectiveness of mRNA BNT162b2 vaccine against SARS-CoV-2 infections and COVID-19 cases, hospitalisations, and deaths following a nationwide vaccination campaign in Israel: an observational study using national surveillance data. Lancet Lond Engl. 2021;397(10287):1819–1829. doi:10.1016/S0140-6736(21)00947-8

9. Walsh EE, Frenck RW, Falsey AR, et al. Safety and Immunogenicity of Two RNA-Based Covid-19 Vaccine Candidates. N Engl J Med. 2020;383(25):2439–2450. doi:10.1056/NEJMoa2027906

10. Ramasamy MN, Minassian AM, Ewer KJ, et al. Safety and immunogenicity of ChAdOx1 nCoV-19 vaccine administered in a prime-boost regimen in young and old adults (COV002): a single-blind, randomised, controlled, phase 2/3 trial. The Lancet. 2020;396(10267):1979–1993. doi:10.1016/S0140-6736(20)32466-1

11. Les chiffres du R.E.I.N. - Agence de la biomédecine. Published July 1, 2021. Accessed July 2, 2021. https://www.agence-biomedecine.fr/Les-chiffres-du-R-E-I-N

