## Supplementary Material for "Vaccination and COVID-19 dynamics in hemodialysis patients: a population-based study in France"

### **Statistical model**

We modeled weekly incidences SARS-CoV-2 severe infections (COVID-19 cases leading to hospital admissions) in maintenance hemodialysis (MHD) patients from same-age incidences in the general population, with the use of hierarchical Bayesian Poisson regressions accounting for spatial autocorrelation. Data was aggregated spatially at the department level (administrative division of France; there are 96 metropolitan departments) and temporally at the week level. Incidences and vaccination coverages were considered by 10-year age classes (25-35, 35-45, 45-55, 55-65, 65-75, 75-85 and > 85 years).

All multivariable models were built from the following pattern:

$$log\left( \frac{E\left( n_{a,d,w} \right)}{e_{a,d,w}} \right)=\alpha+\beta log\left( N_{a,d,w} \right)+\gamma_{a} a+\ldots+\omega_{d}$$

With:

- $E\left( n_{a,d,w} \right)$: expected number of cases in MHD patients for age class $a$ in department $d$ on week $w$ ($n_{a,d,w}$ following a Poisson distribution);
- $e_{a,d,w}$: exposed (at-risk) MHD patients for the same age / department / week;
- $N_{a,d,w}$: number of cases in the general population for the same age / department / week;
- $a, \beta, \gamma_{a}$: fixed effects to be estimated;
- $\omega_{d}$: random effect accounting for spatial autocorrelation in department $d$;
- $\ldots$ : optional predictors with their associated fixed effects.

Spatial random effects were estimated with a covariance structure depending on neighborhood departments from a BYM model (1). Bayesian inference was performed using integrated nested Laplace approximation (2) and weakly informative priors.

Optional predictors were considered to fit the models:

- number of cases in the general population for the same age / department but for the previous week (smoothing parameter);
- total number of cases in the general population for the same week / department;
- epidemic wave (dummy variables to identify epidemic waves 1 to 3);
- vaccination coverage (1^st^ dose) in the general population for the same week / department (either in the same age class or globally);
- vaccination coverage (1^st^ dose) in MHD patients for the same week / department (accurate enough data stratified by age class was not available).

Other regression models (zero-inflated Poisson and negative binomial) were also considered to improve goodness-of-fit.

Three main models were built:

- M_1_, fitted from 1^st^ wave data, was used to compare 2^nd^ wave predictions to reported cases and identify changes in factors associated with incidence in MHD patients from the beginning of the 2^nd^ wave ;
- M_2_, fitted from 1^st^ and 2^nd^ wave data, was used to compare 3^rd^ wave predictions to reported cases and identify changes in factors associated with incidence in MHD patients from the beginning of the 3^rd^ wave;
- M_3_, fitted from 1^st^ to 3^rd^ wave data, was used to identify factors associated with incidence in MHD patients globally.

Predictors were selected to improve goodness-of-fit according to the Watanabe–Akaike information criterion (WAIC).

The following models were selected:

| M_1_: | $log\left( \frac{E\left( n_{a,d,w} \right)}{e_{a,d,w}} \right)=\alpha+\beta log\left( N_{a,d,w} \right)+\beta^{'}log\left( N_{a,d,w-1} \right)+\gamma_{a} a+\omega_{d}$ |
| --- | --- |

| M_2_: | $log\left( \frac{E\left( n_{a,d,w} \right)}{e_{a,d,w}} \right)=\alpha+\beta log\left( N_{a,d,w} \right)+\beta^{'}log\left( N_{a,d,w-1} \right)+\delta k_{w}+\gamma_{a} a+\omega_{d}$ |
| --- | --- |

With $k_{w}$ taking values according to the epidemic wave ($k_{w}$ = 0 if week $w$ falls into the 1^st^ wave and 1 otherwise) and $\delta$ the associated fixed coefficient to be estimated.

| M_3_: | $log\left( \frac{E\left( n_{a,d,w} \right)}{e_{a,d,w}} \right)=\alpha+\beta log\left( N_{a,d,w} \right)+\beta^{'}log\left( N_{a,d,w-1} \right)+\delta k_{w}+\gamma_{a} a+\xi_{0} u_{a,d,w-5}+\xi_{a} a\times u_{a,d,w-5}+\zeta v_{a,d,w-3}+\omega_{d}$ |
| --- | --- |

With $u_{a,d,w}$ and $v_{a,d,w}$ the vaccination coverages in MHD patients and in the general population, respectively, for age class $a$ on week $w$ in department $d$, $\xi_{0}$ and $\zeta$ the associated fixed coefficients and $\xi_{a}$ the coefficient for interaction between $a$ and $u_{a,d,w}$ to be estimated. The 3- and 5-week time lapses were set to account for a humoral response likely to impact incidence.

All analyses were conducted with R statistical software version 4.0. Models were fitted with the INLA package (3).

### **Supplementary table S1 : Observed and predicted number of severe SARS-CoV-2 infections in patients requiring maintenance hemodialysis (MHD)**

|  | **1^st^ wave** | **2^nd^ wave** | **3^rd^ wave** | **Total** |
| --- | --- | --- | --- | --- |
| Observed cases | 1233 | 1447 | 940 | 3620 |
| M_1_ predictions | 1265 (1156, 1397) | 2092 (1888, 2317) | 2021 (1874, 2194) | 5378 (5031, 5868) |
| M_2_ predictions | 1246 (1137, 1377) | 1475 (1371, 1577) | 1457 (1356, 1557) | 4179 (3957, 4382) |
| M_3_ predictions | 1258 (1190, 1333) | 1461 (1386, 1544) | 957 (938, 998) | 3685 (3552, 3816) |

Predicted number of MHD severe infections (infection leading to hospital admissions) with 95% credible intervals from models M_1_ (fitted on 1^st^ wave data, March 11 to June 30, 2020), M_2_ (fitted on 1^st^ and 2^nd^ wave data, up to July 1^st^, 2020) and M_3_ (fitted on all 3 waves, up to April 29, 2021).

### **Supplementary table S2 : Regression Models coefficients**

| **Model M_1_** | |  | |
| --- | --- | --- | --- |
| **Variable** | | | **IRR (95% CI)** |
| - $log\left( N_{a,d,w} \right)$: (log) number of cases in the general population | | | 2.12 (1.90, 2.37) |
| - $log\left( N_{a,d,w-1} \right):$(log) number of cases in the general population the previous week | | | 1.79 (1.59, 2.03) |
| - $a$: age class - 25-35 years - 35-45 years - 45-55 years - 55-65 years - 65-75 years - 75-85 years - > 85 years | | | 4.04 (2.38, 6.51)  2.57 (1.85, 3.52)  1.37 (1.06, 1.76)  1 (reference)  0.85 (0.71, 1.02)  0.87 (0.73, 1.04)  0.90 (0.73, 1.10) |
| **Model M_2_** | | |  |
| **Variable** | | | **IRR (95% CI)** |
| - $log\left( N_{a,d,w} \right)$: (log) number of cases in the general population | | | 1.98 (1.81, 2.16) |
| - $log\left( N_{a,d,w-1} \right):$(log) number of cases in the general population the previous week | | | 1.68 (1.54, 1.84) |
| - $a$: age class - 25-35 years - 35-45 years - 45-55 years - 55-65 years - 65-75 years - 75-85 years - > 85 years | | | 2.82 (1.90, 4.06)  2.17 (1.70, 2.75)  1.26 (1.04, 1.52)  1 (reference)  0.83 (0.74, 0.94)  0.74 (0.66, 0.85)  0.82 (0.72, 0.95) |
| - $k_{w}$: epidemic wave ≥ 2 | | | 0.70 (0.64, 0.76) |
| **Model M_3_** | | |  |
| **Variable** | | | **IRR (95% CI)** |
| - $log\left( N_{a,d,w} \right)$: (log) number of cases in the general population | | | 1.91 (1.76, 2.08) |
| - $log\left( N_{a,d,w-1} \right):$(log) number of cases in the general population the previous week | | | 1.69 (1.56, 1.84) |
| - $a$: age class - 25-35 years - 35-45 years - 45-55 years - 55-65 years - 65-75 years - 75-85 years - > 85 years | | | 2.92 (2.03, 4.09)  2.19 (1.74, 2.73)  1.32 (1.11, 1.56)  1 (reference)  0.88 (0.78, 0.98)  0.78 (0.70, 0.88)  0.86 (0.76, 0.97) |
| - $k_{w}$: epidemic wave ≥ 2 | | | 0.70 (0.64, 0.76) |
| - $v_{a,d,w-3}$: Vaccination coverage in the general population   (per 10% increase) | | | 0.50 (0.40, 0.61) |
| - $u_{a,d,w-5}$: Vaccine exposure in MHD patients | | | 0.37 (0.18, 0.71) |
| - ${a\times u}_{a,d,w-5}$: Age–vaccine interaction - 25-35 years - 35-45 years - 45-55 years - 55-65 years - 65-75 years - 75-85 years - > 85 years | | | 0.00 (0.00, 4.66)  0.04 (0.00, 0.46)  0.55 (0.16, 1.72)  reference  0.94 (0.43, 2.11)  1.74 (0.80, 3.86)  2.30 (0.95, 5.56) |

Coefficients from the Poisson regression models are exponentiated and reported as incidence rate ratios (IRR) with their 95% confidence intervals (CI).

### **References**

1.	Besag J, York J, Mollié A. Bayesian image restoration, with two applications in spatial statistics. Ann Inst Stat Math. 1991 Mar 1;43(1):1–20.

2.	Gomez-Rubio V. Bayesian inference with INLA. CRC Press; 2020. 308 p.

3.	Rue H, Martino S, Chopin N. Approximate Bayesian inference for latent Gaussian models by using integrated nested Laplace approximations. Journal of the Royal Statistical Society: Series B (Statistical Methodology). 2009;71(2):319–92.
